# Real world evidence of acute interstitial lung disease-related hospital admissions infers complex, multifactorial association between social deprivation and 90-day all-cause mortality outcomes: data from the North West of England

**DOI:** 10.64898/2026.03.12.26348240

**Authors:** Laura White, Jonathon Shaw, Bethan Powell, Nyan May Kyi, Alicia Sou, Gareth Hughes, Dilanka Tilakaratne, Conal Hayton, Trishala Raj, Vi Truong, Nashwah Ismail, Nawat Khanijoun, Rebecca Huang, Emma Hardy, Mahzaib Babar, Naayaab Khan, Martin Regan, Oby Okpala, Ragavilasini Suresh, Jerome McIntosh, Amsal Amjad, Mahum Sohail, Zainab Aslam, Amy Gadoud, Timothy Gatheral, Georges NgManKwong

## Abstract

**Background:** Social deprivation impacts chronic disease and acute admission outcomes. In interstitial lung disease (ILD), prior British Thoracic Society registry data for idiopathic pulmonary fibrosis has shown high deprivation was associated with poorer long-term outcomes. However, its impact on acute admissions in ILD is not known.

**Methods:** We undertook a multicentre, retrospective study of ILD-related admissions between 1^st^ January 2017 and 31^st^ December 2019 across 11 hospitals in the North West of England, utilising available real-world data. We determined social deprivation geographically by the 2019 English Indices of Deprivation deciles. The primary outcome was 90-day all-cause mortality.

**Results:** 999 admissions met the inclusion criteria. 327/999 (32.7%) of admissions came from individuals geographically in the most deprived 20%. Across 999 admissions, in unadjusted survival analysis we observed a non-linear relationship between deprivation and 90-day all-cause mortality. In complete case multivariate modelling, deprivation demonstrated borderline significant association with all-cause mortality (HR 1.038, 95% CI 1.00 – 1.077, p = 0.050). However, this effect was lost in pooled analysis using multiple imputation (HR 1.001, 95% CI 0.971 – 1.033, p = 0.928). Male sex and pre-admission long-term oxygen were consistently associated with increased 90-day all-cause mortality across both models. Lower TLCO values were significantly associated with increased 90-day mortality in pooled analysis.

**Conclusion:** We observe a high burden of acute ILD-related hospital admission amongst the most deprived 20%, suggesting geographical deprivation may impact acute healthcare seeking behaviours. Once admitted, the impact of deprivation appears more complex and multifactorial. Further studies which assess geographical and individual-level deprivation are needed to validate our findings.

**Key Messages:** 

**What is already known on the topic?:** The British Thoracic Society idiopathic pulmonary fibrosis registry has previously demonstrated that higher social deprivation is associated with worse long-term outcomes. In other respiratory diseases, social deprivation impacts acute admission patterns and outcomes.

**What this study adds:** To the best of our knowledge, this is the first study examining the relationship between social deprivation and acute ILD-related admission outcomes. This study demonstrates high acute admission burden from the geographically most deprived 20%. Once admitted, the association between geographical social deprivation and mortality outcomes appears complex and multifactorial in our modelling.

**How this may affect research, practice or policy:** This study highlights the acute admission burden from highly deprived communities and the need for additional research to further understand the individual-level and geographical-level deprivation patients with ILD experience. We suggest the need for community outreach to build trust with deprived communities, alongside increasing awareness amongst patients, caregivers and primary care physicians in such communities. Deprivation must remain an important consideration in any new service or intervention to prevent worsening of health inequalities.

## Introduction

Social determinants of health (SDH) describe the nonmedical components of health which influence disease outcomes.[1] Social deprivation is a complex entity which describes one component of this. According to the UK Government, deprivation is defined as an individual who lacks “any kind of resources”, whereas poverty refers to the specific lack of financial resources to meet their needs.[2] Inequalities in health and outcomes in chronic disease are closely associated with SDH. The World Health Organisation (WHO) estimate nonmedical factors contribute to between 30% and 55% of disease outcomes.[3]

In the 2019 English indices of deprivation, seven domains are used to determine an Index of Multiple Deprivation (IMD) for each small locality, related to postcode. These domains include weighted averages of income (22.5%), employment (22.5%), education (13.5%), health and disability (13.5%), crime (9.3%), barriers to housing and services (9.3%), and living environment (9.3%).[4] Local authorities are subsequently ranked and split into ten, determining the ten deciles of deprivation. Importantly, these deciles reflect average geographical deprivation – rather than individual-level deprivation statistics.

Interstitial lung diseases (ILD) represent a heterogenous group of disorders affecting the lung parenchyma. Idiopathic pulmonary fibrosis (IPF) is the archetypal progressive fibrotic form of ILD. Data from the British Thoracic Society (BTS) IPF registry found those from the most deprived social backgrounds had more severe disease at diagnosis and worse long-term outcomes.[5] In ILD more generally, living and working conditions were most frequently associated with health inequalities – and thus overall patient outcomes.[3] The North West of England has the highest rate of deprivation in the UK, with 45% of districts ranked in the most deprived decile located in the region.[6] Given the evidence of health inequality in chronic disease, combined with high levels of social deprivation in the North West, we sought to determine if social deprivation impacts 90-day mortality outcomes from acute ILD-related hospital admissions in this region.

## Methods

### Study Setting and Population

We performed a multicentre retrospective observational study of ILD-related hospital admissions in adults ≥18 years old. Admissions were identified using International Classification of Disease Version 10 coding between 01.01.2017 and 31.12.2019 from 11 NHS secondary and tertiary hospitals in the North West of England (supplementary figure 1). ICD-10 codes of B22.1, D86.0, D86.2, J67.0-67.9, J70.2-70.4, J84.1, J84.8 and J84.9 within the primary diagnosis of admission event triggered review for inclusion in the study. Reasons for exclusion are summarised in supplementary figure 1, including any prior documented dissent for use of data in research.

Baseline admission and patient characteristics data were sourced from coding and manual searching of medical records. Full postcodes were converted to deprivation deciles (DD) as per the 2019 UK Government English Indices of Deprivation data, with a DD of 1 representing the most deprived 10%, and a DD of 10 the least deprived 10%.[2] Admission events meeting criteria were subsequently grouped into quintiles, with quintile 1 representing the least deprived (DD’s 9 and 10) and quintile 5 representing the most deprived (DD’s 1 and 2). Primary outcome was number of days from start of admission to death.

### Patient and Public Involvement

Following the development of the study protocol, the concept and planned design was presented to a group of expert patients, the Research Champions through Action for Pulmonary Fibrosis, prior to IRAS submission. They confirmed acceptability of the proposed design and assisted with planning of a dissemination strategy, including radio and newsletter appearances following publication. Given the retrospective data design of the study, no further patient or public involvement was undertaken.

The Action for Pulmonary Fibrosis Research Champions are a group of expert patients who are involved in charitable and research activities related to ILD. While it was beneficial to discuss the proposed study with this group, further details on their socioeconomic status (SES) were not requested nor recorded. In retrospect, given the significant proportion of acute admissions from the most deprived 20% of the population, this would have been beneficial to understanding the relevance for the population these results impact. It is therefore an important limitation of our patient and public involvement that future studies should look to address. This likely includes going out into deprived communities, perhaps away from structured support group settings, to gather this input.

### Data Collection and Variables

All data was sourced from coding and manual searching of medical records by site-specific clinicians. All admissions meeting inclusion criteria underwent manual review of medical notes. Data, where available, was collected on patient demographics, co-morbidities (converted to Charlson Comorbidity Index (CCI)[7]) and pre-existing ILD diagnoses. Underlying ILD diagnosis was obtained from manual searching of medical records, using multidisciplinary team (MDT) documentation, radiology and clinic letters where available. During data collection, we observed a group of patients without an MDT-confirmed diagnosis, but in whom evidence of fibrotic ILD was present. This group was labelled “pulmonary fibrosis as a diagnostic label” (PFD), reflecting real-world access to ILD MDT diagnosis. Additional ILD-specific data including antifibrotic use, pre-admission oxygen use and pre-admission lung function testing was recorded where available.

Admission-relevant data included mortality outcomes and at-admission biochemical investigations (white cell count and differential, C-reactive protein (CRP)). As part of the manual search of medical records, site-specific clinicians assessed the admission reason using information available from the clinical narrative, biochemical and radiological investigations. For the purposes of this study, an acute exacerbation of interstitial lung disease (AEILD) event was defined clinically by <30-day clinical deterioration, not secondary to cardiac failure, thromboembolic disease (TED) or pneumothorax event. Whether AEILD criteria was met was at the discretion of the site-specific clinician. If it was not felt to be met, they were deemed to have an ‘other’ ILD-related hospital admission. Other ILD-related admission reasons included disease progression, pulmonary embolism, pulmonary hypertension and pneumothorax but were not recorded as discrete variables.

### Statistical Analysis

Statistical analysis was undertaken using SPSS v31.0 and Python v3.9.7. Continuous data were assessed for normality with Shapiro-Wilk test. Statistical comparison between quintiles was undertaken by one-way ANOVA or Kruskal-Wallis H, chi-squared or Fisher’s exact test as appropriate, with post-hoc analysis using Dunn’s test for continuous variables and standardised residuals for categorical variables, with Bonferroni correction. Time-to-event survival analysis was undertaken by Kaplan-Meier with log-rank comparison.

Hazard ratios were initially estimated using a complete case multivariate Cox proportional hazards model with the ENTER method – whereby all variables are entered into the model simultaneously. Variable selection for the model was based on clinical relevance and prior evidence in mortality outcomes, rather than data-driven stepwise methods, to minimise model overfitting. Demographic characteristics (including age, gender, ethnicity) were included to assess the impact of non-modifiable risk factors, while clinical and inflammatory variables were chosen for their known or hypothesised relevance to mortality risk in ILD populations.[8–10] Variables with high proportions of missing data were not included (with a threshold of ≥15% missing data used) for complete case analysis. As a result, forced vital capacity (FVC, 387/999 or 38.7% missing), transfer factor for carbon monoxide (TLCO, 596/999 or 59.6% missing) and oxygen (L/min) at admission (679/999, or 67.9% missing) were not included in the model due to high levels of data missingness. While rates of antifibrotic medication use were statistically significantly different between the quintiles (p = 0.002; table 1), there was an overall low rate of antifibrotic use. As such, this was not included in the multivariate modelling due to the risk of model over-fitting and inaccurate estimates of mortality association due to small numbers. The proportional hazards assumption was checked using Schoenfeld residuals.

**Table 1:**
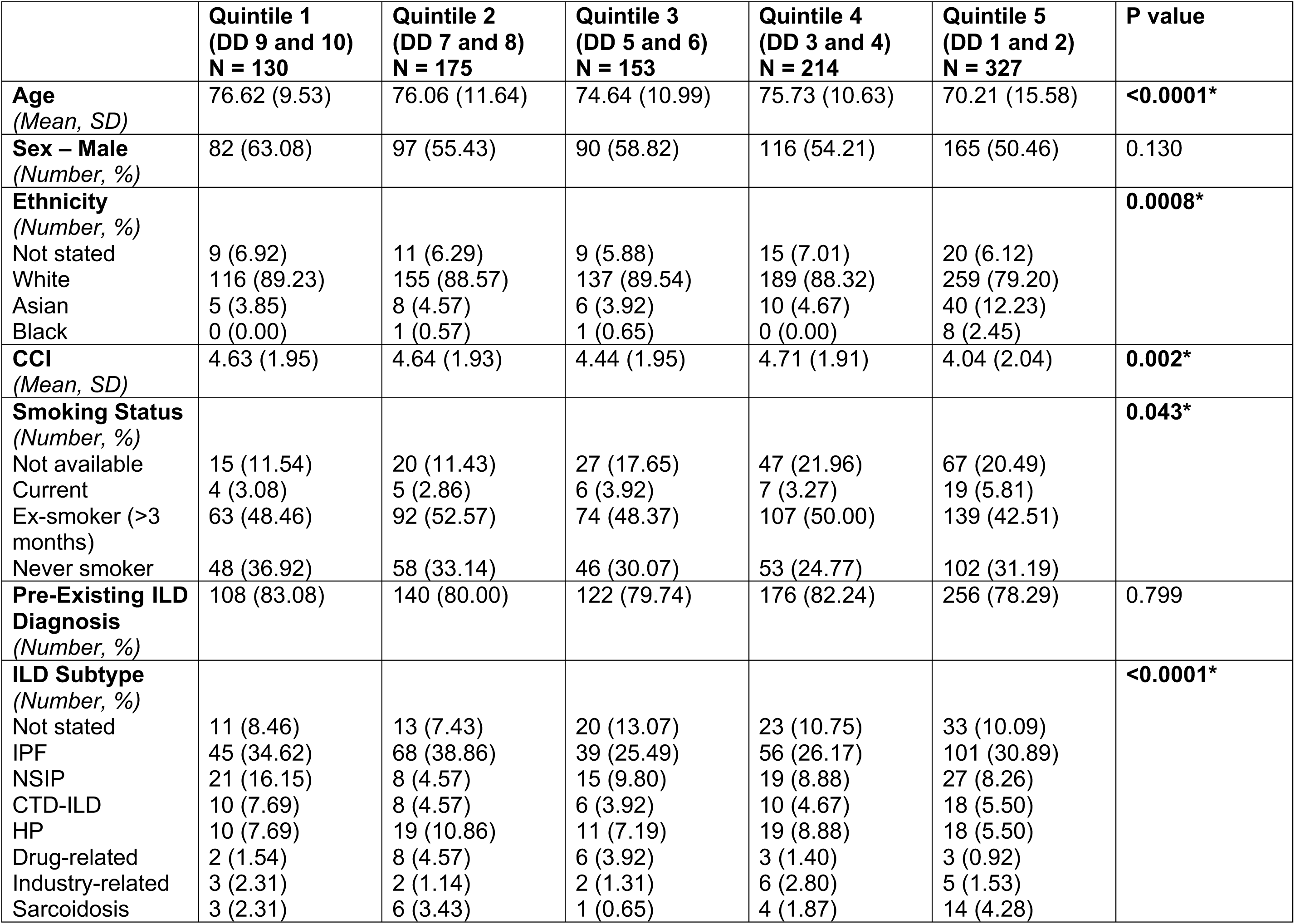
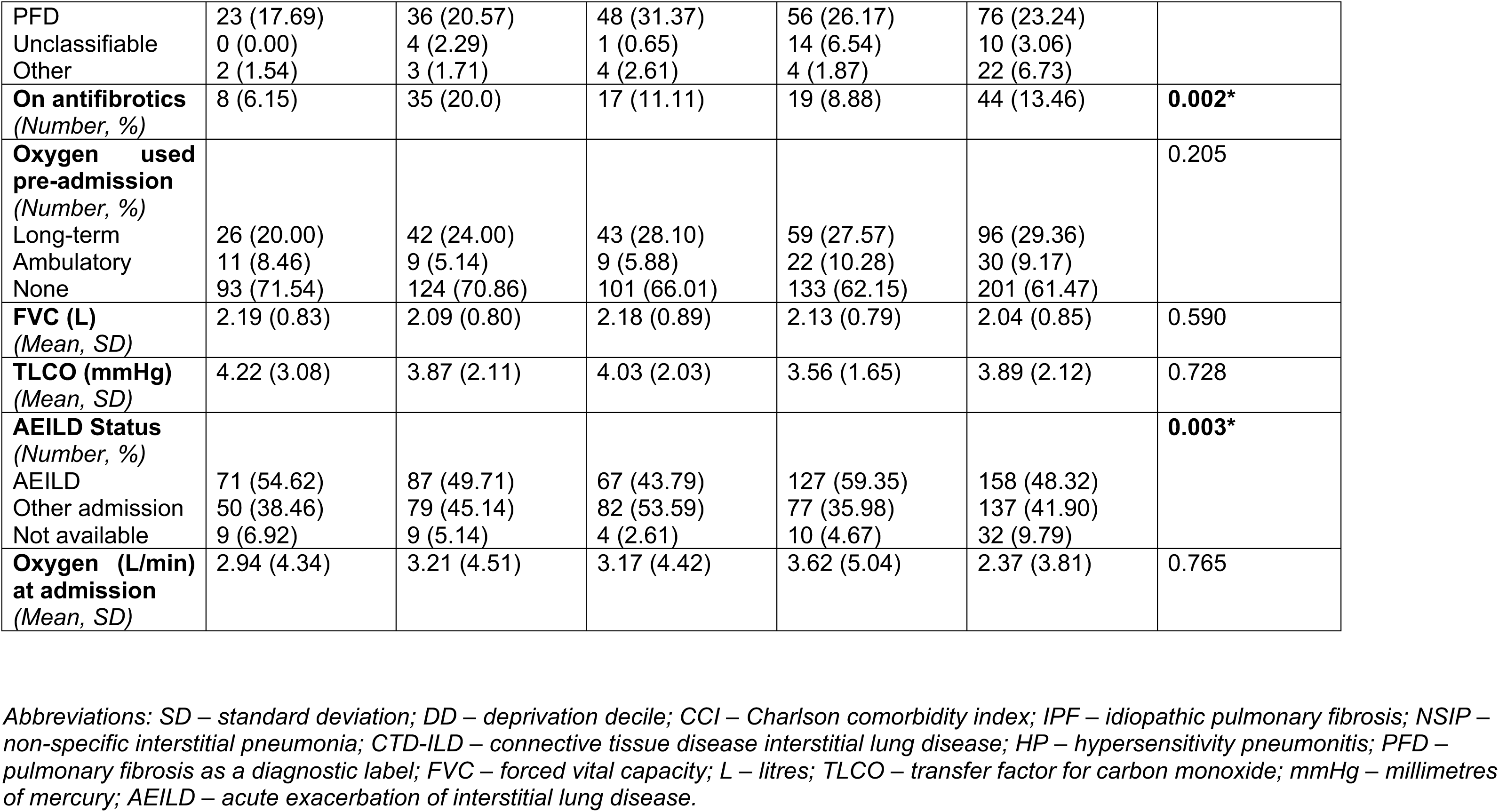
Full descriptive baseline characteristics of each deprivation quintile, based on the 2019 English Indices of Deprivation. Quintile 1 represents the least deprived 20%, equivalent to deprivation deciles 9 and 10, and quintile 5 the most deprived 20%, equivalent to deprivation deciles 1 and 2. Continuous variables were described using mean (standard deviation) and subsequently tested for normality with the Shapiro-Wilk test. Quintiles were subsequently compared using ANOVA or Kruskal-Wallis, dependent on normality. Categorical variables were described using number frequency (percentage) and compared using chi-squared or Fisher’s exact test, dependent on expected values greater or less than five. A global P value is reported. Statistically significant values are marked in bold and with an asterixis (*). Additional note: post-hoc pairwise comparison analysis for age and CCI using Dunn’s test with Bonferroni correction available as supplementary table 1. Post-hoc analysis for categorical variables using standardised residuals and pairwise comparison with Bonferroni correction is available as supplementary table 2.

|  | **Quintile 1**<br>**(DD 9 and 10)**<br>**N = 130** | **Quintile 2**<br>**(DD 7 and 8)**<br>**N = 175** | **Quintile 3**<br>**(DD 5 and 6)**<br>**N = 153** | **Quintile 4**<br>**(DD 3 and 4)**<br>**N = 214** | **Quintile 5**<br>**(DD 1 and 2)**<br>**N = 327** | **P value** |
| --- | --- | --- | --- | --- | --- | --- |
| **Age**<br>(Mean, SD) | 76.62 (9.53) | 76.06 (11.64) | 74.64 (10.99) | 75.73 (10.63) | 70.21 (15.58) | **<0.0001\*** |
| **Sex – Male**<br>(Number, %) | 82 (63.08) | 97 (55.43) | 90 (58.82) | 116 (54.21) | 165 (50.46) | 0.130 |
| **Ethnicity**<br>(Number, %) |  |  |  |  |  | **0.0008\*** |
| Not stated | 9 (6.92) | 11 (6.29) | 9 (5.88) | 15 (7.01) | 20 (6.12) |  |
| White | 116 (89.23) | 155 (88.57) | 137 (89.54) | 189 (88.32) | 259 (79.20) |  |
| Asian | 5 (3.85) | 8 (4.57) | 6 (3.92) | 10 (4.67) | 40 (12.23) |  |
| Black | 0 (0.00) | 1 (0.57) | 1 (0.65) | 0 (0.00) | 8 (2.45) |  |
| **CCI**<br>(Mean, SD) | 4.63 (1.95) | 4.64 (1.93) | 4.44 (1.95) | 4.71 (1.91) | 4.04 (2.04) | **0.002\*** |
| **Smoking Status**<br>(Number, %) |  |  |  |  |  | **0.043\*** |
| Not available | 15 (11.54) | 20 (11.43) | 27 (17.65) | 47 (21.96) | 67 (20.49) |  |
| Current | 4 (3.08) | 5 (2.86) | 6 (3.92) | 7 (3.27) | 19 (5.81) |  |
| Ex-smoker (>3 months) | 63 (48.46) | 92 (52.57) | 74 (48.37) | 107 (50.00) | 139 (42.51) |  |
| Never smoker | 48 (36.92) | 58 (33.14) | 46 (30.07) | 53 (24.77) | 102 (31.19) |  |
| **Pre-Existing ILD Diagnosis**<br>(Number, %) | 108 (83.08) | 140 (80.00) | 122 (79.74) | 176 (82.24) | 256 (78.29) | 0.799 |
| **ILD Subtype**<br>(Number, %) |  |  |  |  |  | **<0.0001\*** |
| Not stated | 11 (8.46) | 13 (7.43) | 20 (13.07) | 23 (10.75) | 33 (10.09) |  |
| IPF | 45 (34.62) | 68 (38.86) | 39 (25.49) | 56 (26.17) | 101 (30.89) |  |
| NSIP | 21 (16.15) | 8 (4.57) | 15 (9.80) | 19 (8.88) | 27 (8.26) |  |
| CTD-ILD | 10 (7.69) | 8 (4.57) | 6 (3.92) | 10 (4.67) | 18 (5.50) |  |
| HP | 10 (7.69) | 19 (10.86) | 11 (7.19) | 19 (8.88) | 18 (5.50) |  |
| Drug-related | 2 (1.54) | 8 (4.57) | 6 (3.92) | 3 (1.40) | 3 (0.92) |  |
| Industry-related | 3 (2.31) | 2 (1.14) | 2 (1.31) | 6 (2.80) | 5 (1.53) |  |
| Sarcoidosis | 3 (2.31) | 6 (3.43) | 1 (0.65) | 4 (1.87) | 14 (4.28) |  |
| PFD | 23 (17.69) | 36 (20.57) | 48 (31.37) | 56 (26.17) | 76 (23.24) |  |
| Unclassifiable | 0 (0.00) | 4 (2.29) | 1 (0.65) | 14 (6.54) | 10 (3.06) |  |
| Other | 2 (1.54) | 3 (1.71) | 4 (2.61) | 4 (1.87) | 22 (6.73) |  |
| **On antifibrotics**<br>(Number, %) | 8 (6.15) | 35 (20.0) | 17 (11.11) | 19 (8.88) | 44 (13.46) | **0.002\*** |
| **Oxygen used pre-admission**<br>(Number, %) |  |  |  |  |  | 0.205 |
| Long-term | 26 (20.00) | 42 (24.00) | 43 (28.10) | 59 (27.57) | 96 (29.36) |  |
| Ambulatory | 11 (8.46) | 9 (5.14) | 9 (5.88) | 22 (10.28) | 30 (9.17) |  |
| None | 93 (71.54) | 124 (70.86) | 101 (66.01) | 133 (62.15) | 201 (61.47) |  |
| **FVC (L)**<br>(Mean, SD) | 2.19 (0.83) | 2.09 (0.80) | 2.18 (0.89) | 2.13 (0.79) | 2.04 (0.85) | 0.590 |
| **TLCO (mmHg)**<br>(Mean, SD) | 4.22 (3.08) | 3.87 (2.11) | 4.03 (2.03) | 3.56 (1.65) | 3.89 (2.12) | 0.728 |
| **AEILD Status**<br>(Number, %) |  |  |  |  |  | **0.003\*** |
| AEILD | 71 (54.62) | 87 (49.71) | 67 (43.79) | 127 (59.35) | 158 (48.32) |  |
| Other admission | 50 (38.46) | 79 (45.14) | 82 (53.59) | 77 (35.98) | 137 (41.90) |  |
| Not available | 9 (6.92) | 9 (5.14) | 4 (2.61) | 10 (4.67) | 32 (9.79) |  |
| **Oxygen (L/min) at admission**<br>(Mean, SD) | 2.94 (4.34) | 3.21 (4.51) | 3.17 (4.42) | 3.62 (5.04) | 2.37 (3.81) | 0.765 |

Abbreviations: SD – standard deviation; DD – deprivation decile; CCI – Charlson comorbidity index; IPF – idiopathic pulmonary fibrosis; NSIP – non-specific interstitial pneumonia; CTD-ILD – connective tissue disease interstitial lung disease; HP – hypersensitivity pneumonitis; PFD – pulmonary fibrosis as a diagnostic label; FVC – forced vital capacity; L – litres; TLCO – transfer factor for carbon monoxide; mmHg – millimetres of mercury; AEILD – acute exacerbation of interstitial lung disease.

Due to high levels of data missingness for FVC, TLCO and oxygen (L/min) at admission, an assessment of missingness was undertaken. Logistic regression was used to compare data missingness for FVC, TLCO and oxygen (L/min) at admission with age, CCI, deprivation decile and 90-day mortality outcomes. Associations were observed (supplementary table 4) suggesting missingness may be consistent with a missing-at-random (MAR) mechanism. Subsequent multiple imputation modelling for FVC, TLCO and oxygen (L/min) at admission was undertaken. Multiple imputation using chained equations (MICE) generated 20 imputed datasets. Updated Cox proportional hazards models using the ENTER method were fitted across the imputed datasets and pooled using Rubin’s rules.[11] The imputation model included all variables used in the complete case analysis model. The pooled results are reported. A comparison of hazard ratios (HR) output between complete case analysis and pooled analysis was also undertaken.

Finally, due to significant age differences observed between quintiles in both initial Kruskal-Wallis H analysis (table 1) and in post-hoc pairwise comparison (supplementary table 1), sub-analysis to understand the interaction between age and deprivation was undertaken. An interaction analysis using an age x deprivation term was completed on the pooled analysis model. A p value of <0.05 was considered statistically significant.

## Results

3451 admissions were identified through primary ICD-10 codes relevant to the study. 2452/3451 (71.05%) were excluded, with the predominant reason being attendance at day case procedures (1984/2449, supplementary figure 1). Other reasons for exclusion included transplant work-up attendances (158/2449), inability to access medical record relevant to admission (118/2449), non-ILD related admission (118/2449) and incorrect coding (67/2449, supplementary figure 1). Three admissions were excluded due to absence of postcode to calculate deprivation score, leaving 999 records included in data analysis. Quintile 5, representing the most deprived 20% (DD’s 1 and 2), accounted for 327/999 (32.7%) of admission events.

### Baseline Characteristics

Complete descriptions of the baseline characteristics for each quintile cohort are described in table 1. Several variables showed statistically significant differences at initial data interrogation: age, ethnicity, CCI, smoking status, ILD subtype, antifibrotic use and AEILD status.

Post-hoc analysis supported greater understanding of the differences between quintiles. Pairwise comparison of age demonstrated that quintile 5 was the main outlier, with a statistically significant younger cohort on average when compared to quintile 1 (p = 0.0006), 2 (p <0.0001) and 4 (p = 0.003, supplementary table 1). The difference in co-morbidity burden (as per CCI values) was observed between quintiles 2 and 5 (p = 0.015), and 4 and 5 (p = 0.003), in post-hoc pairwise comparison – with quintile 5 demonstrating the lowest co-morbidity burden using this index (supplementary table 1).

Categorical variables underwent post-hoc analysis interrogation using standardised residuals. Full results of residuals, raw P values and Bonferroni correct P values for smoking status, ethnicity, ILD subtype, antifibrotics and AEILD status are available in supplementary table 2. During an initial assessment, smoking status demonstrated a statistically significant chi-squared value, however this effect was not demonstrated in post-hoc analysis. Similar patterns were observed for ILD subtype, antifibrotic use, ethnicity and ILD status. Of note however, prior to Bonferroni correction, analysis suggested a trend to increased numbers of NSIP sub-type within quintile 1 (raw p value = 0.007, supplementary table 2), versus reduced numbers of NSIP within quintile 2 (raw p value = 0.050, supplementary table 2) – but statistical significance was not observed with correction. Quintile 5 demonstrated a pattern of more patients with an “other” ILD subtype (raw P value = 0.002, supplementary table 2), inclusive of several ILD subtypes, but this did not remain significant at the point of Bonferroni correction.

### Primary Outcome: 90-day All-Cause Mortality

The overall 90-day mortality across all quintiles was 39.1%. Time-to-event analysis demonstrated overall statistically significant difference in 90-day all-cause mortality between the deprivation quintiles observed (p <0.002, figure 2) with subsequent pairwise comparison revealing significant difference in survival between quintile 2 and all other quintiles (2 vs. 1 p = 0.036; 2 vs. 3 p = 0.022; 2 vs. 4 p = 0.003; 2 vs. 5 p = 0.0001, figure 2).

### Secondary Outcomes: Multivariate Modelling of 90-day all-cause Mortality

Eleven variables were included in a multivariate model for 90-day all-cause mortality with complete case analysis (table 2). Prior to completing, the proportional hazards assumption was checked. There was no evidence of widespread violation. Minor violations were suggested for CCI and one ILD subtype, but no substantial overall violations were identified. Key variables associated with increased risk of mortality in this model included male gender (HR 1.343, 95% CI 1.070 -1.687, p = 0.011; table 2), long-term oxygen therapy (HR 1.931, 95% CI 1.528 – 2.440, p = <0.0001, table 2) and increased neutrophil count (HR 1.083, 95% CI 1.052 – 1.115, p <0.0001, table 2). A higher deprivation decile (representing lower deprivation) showed a borderline association with increased mortality (HR of 1.038, 95% CI 1.000 – 1.077, p = 0.050).

**Table 2:** Full results of multivariate cox regression analysis of 90-day all-cause mortality associated with interstitial lung disease-related hospital admissions, using complete case analysis. Statistically significant values are marked in bold and with an asterixis (*).

|  | HR | Lower 95% CI | Upper 95% CI | P value |
| --- | --- | --- | --- | --- |
| **Age** | 1.027 | 1.013 | 1.041 | 0.0002 |
| **Sex** |  |  |  |  |
| Female | Reference | Reference | Reference | Reference |
| Male | 1.343 | 1.070 | 1.687 | **0.011\*** |
| **Ethnicity** |  |  |  |  |
| White | Reference | Reference | Reference | Reference |
| Asian | 0.636 | 0.389 | 1.039 | 0.071 |
| Black | 1.739 | 0.511 | 5.920 | 0.376 |
| Not stated | 1.537 | 1.013 | 2.332 | **0.043\*** |
| **CCI** | 1.012 | 0.941 | 1.087 | 0.752 |
| **Deprivation Decile** | 1.038 | 1.000 | 1.077 | 0.050 |
| **ILD Subtype** |  |  |  |  |
| Not stated | Reference | Reference | Reference | Reference |
| IPF | 1.136 | 0.722 | 1.785 | 0.582 |
| NSIP | 0.547 | 0.203 | 1.473 | 0.232 |
| CTD-ILD | 1.260 | 0.739 | 2.146 | 0.396 |
| HP | 0.652 | 0.329 | 1.295 | 0.222 |
| Drug-related | 0.256 | 0.121 | 0.545 | 0.0004 |
| Industry-related | 0.781 | 0.295 | 2.070 | 0.620 |
| Sarcoidosis | 0.542 | 0.185 | 1.587 | 0.264 |
| PFD | 1.028 | 0.425 | 2.491 | 0.950 |
| Unclassifiable | 0.883 | 0.549 | 1.420 | 0.607 |
| Other | 1.310 | 0.446 | 2.625 | 0.654 |
| **Oxygen** |  |  |  |  |
| None | Reference | Reference | Reference | Reference |
| Long-term | 1.931 | 1.528 | 2.440 | <0.0001 |
| Ambulatory | 1.358 | 0.922 | 1.999 | 0.121 |
| **AEILD Status** |  |  |  |  |
| Insufficient information | Reference | Reference | Reference | Reference |
| AEILD | 1.325 | 0.785 | 2.234 | 0.292 |
| Other ILD-related | 0.904 | 0.527 | 1.550 | 0.713 |
| admission |  |  |  |  |
| **Neutrophils** | 1.083 | 1.052 | 1.115 | <0.0001 |
| **Monocytes** | 0.876 | 0.682 | 1.126 | 0.301 |
| **CRP** | 1.002 | 1.001 | 1.004 | 0.004 |

*Abbreviations: HR – hazard ratio; CI – confidence interval; ILD – interstitial lung disease; CCI – Charlson comorbidity index; IPF – idiopathic pulmonary fibrosis; NSIP – non-specific interstitial pneumonia; CTD-ILD – connective tissue disease interstitial lung disease; HP – hypersensitivity pneumonitis; PFD – pulmonary fibrosis as a diagnostic label; AEILD – acute exacerbation of interstitial lung disease; CRP – C-reactive protein.*

Given the high level of missing data for potentially key variables (FVC, TLCO and oxygen (L/min) at admission), repeat analysis using pooled data from MICE was undertaken to assess the impact on mortality association. Table 3 summarises results from the pooled model. Of note, male gender (HR 1.518, p <0.0001, table 3) and long-term oxygen therapy (HR 2.770, p <0.0001, table 3) remained strongly associated with increased risk of mortality. Lower TLCO values were significantly associated with increased risk of mortality in this model (HR 0.890, 95% CI 0.841 – 0.941, p <0.0001, table 3). A direct comparison of the models is available in table 4.

**Table 3:** Full results of multivariate cox regression analysis of 90-day all-cause mortality associated with interstitial lung disease-related hospital admissions, using multiple imputation modelling for FVC, TLCO and oxygen required at admission (litres). Statistically significant values are marked in bold and with an asterixis (*).

|  | HR | Lower 95% CI | Upper 95% CI | P value |
| --- | --- | --- | --- | --- |
| **Age** | 1.016 | 1.005 | 1.027 | 0.003 |
| **Sex** |  |  |  |  |
| Female | Reference | Reference | Reference | Reference |
| Male | 1.518 | 1.244 | 1.852 | **<0.0001\*** |
| **Ethnicity** |  |  |  |  |
| White | Reference | Reference | Reference | Reference |
| Asian | 0.914 | 0.647 | 1.292 | 0.611 |
| Black | 1.060 | 0.453 | 2.478 | 0.893 |
| Not stated | 0.842 | 0.581 | 1.220 | 0.364 |
| **CCI** | 1.032 | 0.965 | 1.104 | 0.354 |
| **Deprivation Decile** | 1.001 | 0.971 | 1.033 | 0.924 |
| **ILD Subtype** |  |  |  |  |
| Not stated | Reference | Reference | Reference | Reference |
| IPF | 1.285 | 0.921 | 1.794 | 0.140 |
| NSIP | 1.050 | 0.698 | 1.578 | 0.816 |
| CTD-ILD | 0.999 | 0.654 | 1.527 | 0.998 |
| HP | 0.948 | 0.653 | 1.376 | 0.779 |
| Drug-related | 0.634 | 0.360 | 1.118 | 0.116 |
| Industry-related | 1.180 | 0.617 | 2.255 | 0.617 |
| Sarcoidosis | 1.042 | 0.604 | 1.800 | 0.883 |
| PFD | 1.597 | 1.164 | 2.193 | **0.004\*** |
| Unclassifiable | 1.062 | 0.578 | 1.952 | 0.846 |
| Other | 1.245 | 0.771 | 2.001 | 0.369 |
| **Oxygen** |  |  |  |  |
| None | Reference | Reference | Reference | Reference |
| Long-term | 2.770 | 2.113 | 3.633 | **<0.0001\*** |
| Ambulatory | 2.020 | 1.448 | 2.818 | **<0.0001\*** |
| **AEILD Status** |  |  |  |  |
| Insufficient information | Reference | Reference | Reference | Reference |
| AEILD | 1.340 | 0.951 | 1.890 | 0.094 |
| Other ILD-related admission | 1.472 | 1.060 | 2.046 | **0.021\*** |
| **Neutrophils** | 1.028 | 0.995 | 1.062 | 0.092 |
| **Monocytes** | 1.063 | 0.980 | 1.153 | 0.143 |
| **CRP** | 0.998 | 0.996 | 0.999 | **0.028\*** |
| **FVC (L)** | 1.001 | 0.991 | 1.022 | 0.405 |
| **TLCO (mmHg)** | 0.890 | 0.841 | 0.941 | **<0.0001\*** |
| **Oxygen (L/min) at admission** | 0.979 | 0.936 | 1.024 | 0.349 |

*Abbreviations: HR – hazard ratio; CI – confidence interval; ILD – interstitial lung disease; CCI – Charlson comorbidity index; IPF – idiopathic pulmonary fibrosis; NSIP – non-specific interstitial pneumonia; CTD-ILD – connective tissue disease interstitial lung disease; HP – hypersensitivity pneumonitis; PFD – pulmonary fibrosis as a diagnostic label; AEILD – acute exacerbation of interstitial lung disease; CRP – C-reactive protein; FVC – forced vital capacity; L – litres; TLCO – transfer factor of the lung for carbon monoxide; L/min – litres per minute.*

**Table 4:**
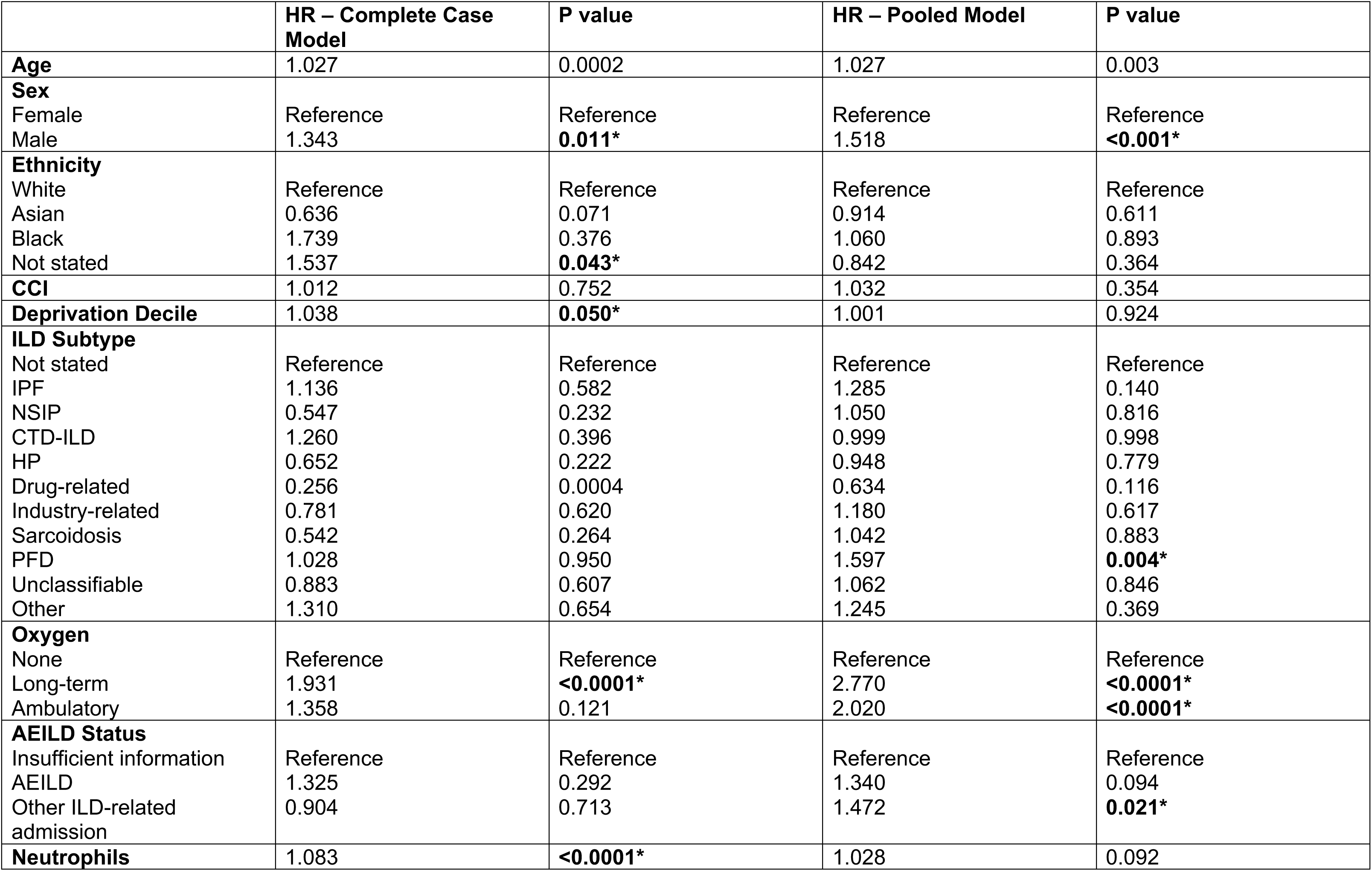
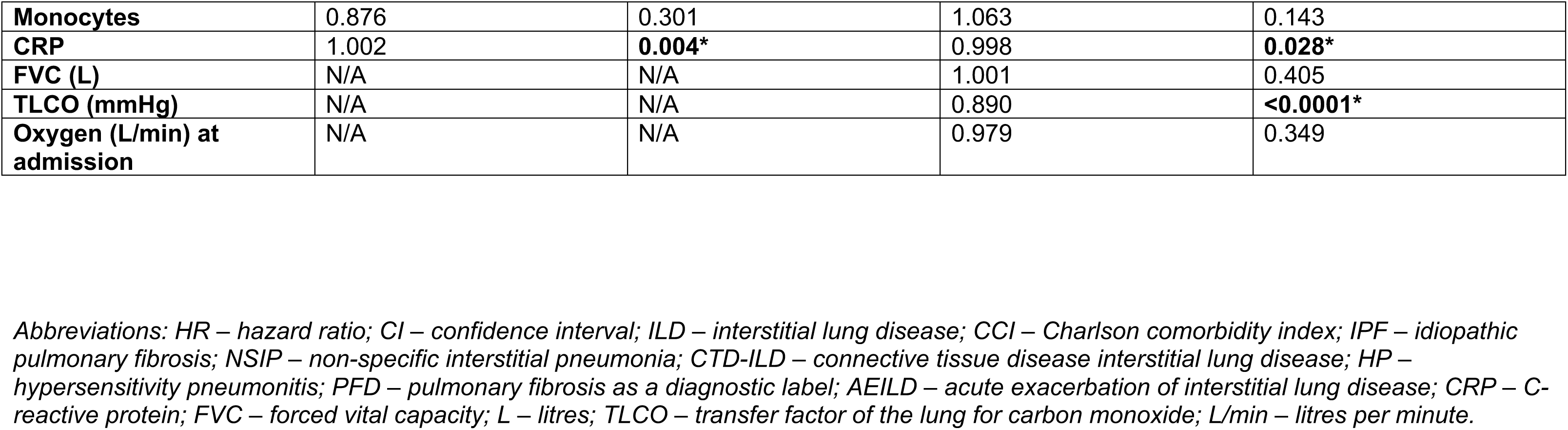
Direct comparison of hazard ratio values between complete case and multiple imputation cox regression modelling. Statistically significant values are marked in bold and with an asterixis (*).

Given the significant age difference observed, multivariate modelling with the pooled data was re-run with an age x deprivation term. In this model, age x deprivation was not significantly associated with increased or reduced risk of mortality (HR 0.999, 95% CI 0.996 – 1.001, p = 0.258, supplementary table 5). The full results of this model are available within supplementary table 5.

## Discussion

### Summary of Results

This retrospective real-world dataset infers a potentially complex, multifactorial association between social deprivation and 90-day all-cause mortality outcomes from acute ILD-related hospital admissions. Of note, we observe high rates of admission amongst the geographically most deprived 20% - with 32.7% (327/999) of acute ILD-related admissions in our dataset from this group.

When considering mortality outcomes, unadjusted survival analysis demonstrates a non-linear relationship between mortality outcomes and social deprivation – with the key difference observed in pairwise comparison between quintile 2 and quintile 5. 90-day all-cause mortality in quintile 2 was 52.3% vs. 33.5% in quintile 5 (p = 0.0001, figure 2). When taken into a complete case multivariate model with eleven variables, lower deprivation demonstrates a borderline increased risk of mortality (HR 1.038, 95% CI 1.000 – 1.077, p = 0.050, table 2). However, this borderline significance is lost in pooled analysis where values for FVC, TLCO and oxygen (L/min) at admission are imputed (HR 1.001, 95% CI 0.971 – 1.033, p = 0.924, table 3). In such multivariate modelling, male gender and pre-admission long-term oxygen therapy remain consistently statistically significantly associated with increased mortality (table 4). In pooled analysis modelling, the significant association of lower TLCO values with increased mortality is demonstrated (HR 0.890, p <0.0001, table 3 and 4).

When comparing across quintiles, significant differences in baseline characteristics were noted. Of particular interest was age, with quintile 5 (the most deprived 20%) demonstrating a statistically significantly reduced age compared to other quintiles, even in post-hoc pairwise comparisons: quintile 1 vs. 5 p = 0.0006; quintile 2 vs. 5 p <0.0001; quintile 3 vs. 5 p = 0.161; quintile 4 vs. 5 p = 0.003 (table 1 and supplementary table 1). Given this, we assessed for the potential of age differences contributing to the pattern observed above. In multivariate modelling, using pooled analysis, an age x deprivation term did not show statistically significant association with mortality (HR 0.999, 95% CI 0.996 – 1.001, p = 0.258, supplementary table 5). Including this term did increase the numerical HR observed for deprivation in multivariate modelling (HR 1.110) but this did not meet statistically significant thresholds (p = 0.256, supplementary table 5).

### Study Strengths and Limitations

A key strength of this study is its multicentre design, encompassing secondary and tertiary hospitals from across the North West of England region. However, there are several important limitations of this study to consider.

This study timeframe (2017 – 2019) was chosen to remove COVID-19 as a potential confounder, given its impact amongst higher deprivation populations.[12] But, we recognise the treatment landscape of ILD has changed significantly since the 2017-2019 period. This is particularly pertinent for the use of antifibrotics – which became much more widespread following the results of the INBUILD study[13] and change in UK NICE guidelines. There have also been significant changes to care pathways in UK ILD services which may impact the results we have observed.[14] Given the small numbers of patients receiving antifibrotics within our cohort, we did not include this variable in our modelling. This is a significant limitation which may impact the relevance of its findings to current populations. Future studies should seek to gather data prospectively to build an accurate picture of the impact of antifibrotics on outcomes – especially within the context of deprivation.

Retrospective data risks recall and misclassification bias – and there were significant challenges with missing data in this dataset (supplementary table 2). As a result, MICE was required to infer the impact of FVC, TLCO and oxygen (L/min) at admission on 90-day mortality outcomes. It also meant we were unable to reliably include the GAP index in our multivariate modelling. Given this, our results may not reflect true outcomes and future studies should aim to gather data prospectively so that more complete datasets can be achieved. Additionally, there is the potential for misclassification bias given AEILD status was based upon the opinion of the site-specific data collector. Inter-rate reliability was not assessed, and thus consistency in diagnosis may be impacted. Admission coding adds to this challenge. Given the potential for incorrect coding, it is unlikely all cases from across this period have been identified – which may have impacted the deprivation skew we observed.

A further limitation is the baseline deprivation within the North West of England population. We recognise that this region has, comparatively, high levels of deprivation and as such our dataset is skewed to including patents from areas of high deprivation. This may limit the generalisability of our findings. However, geographical deprivation does not always equate to individual level deprivation. In future prospective studies, researchers should look to include more detailed individual-level deprivation markers.

### Findings in Context

Our findings are contrary to previous observations in chronic forms of ILD[5] – raising important questions about why this has been observed. To the best of our knowledge, this is the first-time social deprivation has been studied in the setting of acute ILD-related admissions. Acute hospitalisations, including acute exacerbations, are well-recognised to be significant events in the trajectory of ILD.[15–16] However, our findings suggest a potentially complex, multifactorial association between social deprivation and 90-day all-cause mortality in acute ILD-related hospital admissions.

Our unadjusted survival analysis revealed a non-linear relationship between deprivation and mortality outcomes, with those experiencing low-intermediate deprivation (DD’s 7 and 8, quintile 2) demonstrating a significantly worse survival compared to many other deprivation quintiles (figure 2). There are several potential explanations for this. One is the difference in age and comorbidity burden, particularly between quintile 2 and 5, which remains significant in post-hoc analysis (supplementary table 1). In other conditions, such as community-acquired pneumonia, age and comorbidities can interact to worsen mortality outcomes[17] – and thus there is potential these factors play significant roles. However, in multivariate modelling, we did not observe age or comorbidities (reported as CCI) as significant factors associated with mortality (tables 2, 3 and 4) – even when a specific age x deprivation term is developed (supplementary table 5).

Other potential explanations for this non-linear relationship may include more advanced pre-existing ILD prior to the admission event. Differences in TLCO, especially disproportionally lower TLCOs, may infer co-existing pulmonary arterial hypertension (PAH). Co-existing PAH has previously been associated with poorer mortality outcomes.[18] Such patterns of disease severity in our cohort may occur due to chance, or due to true differences between the cohorts. Using available data, we did not observe a statistically significant difference across the quintiles in mean FVC values or mean TLCO values (table 1). This must be interpreted with high caution given the high rates of missing data – meaning absolute conclusions cannot be drawn. Prospective studies should be undertaken to re-assess this possibility, especially given the strong signal in multivariate modelling suggesting lower TLCO values may be associated with mortality outcomes (table 3), consistent with prior findings[19–20].

A final explanation for these findings is that DD is not representative of true individual-level deprivation. The UK 2019 English Indices of deprivation use averages of seven deprivation factors within an area, resulting in postcodes being representative of geographical level deprivation. IMD’s have been shown to be poor proxy’s for individual income, especially amongst groups with poor health.[21] In COVID-19, geographical deprivation as a marker of individual deprivation may have resulted in inaccurate assessments.[22] As such, our marker of deprivation may not reflect the true individual-level deprivation score – resulting in an over- or under-estimation of deprivation and skewing of our quintiles subsequently. Future studies should look to include additional markers of individual deprivation, such as: dental health[23], educational attainment and air pollution.[24]

As such, we hypothesise the non-linear relationship observed in unadjusted analysis may be secondary to multiple, likely interacting factors, that are not fully explained in our dataset. Future studies are needed to first understand if this pattern is observed in repeat datasets.

### Admission Patterns: The Impact of Social Deprivation

A key finding of our retrospective dataset is the high admission burden from the most deprived 20%. It is well recognised that emergency attendances and admission rates are higher amongst the most deprived[25], reflecting the challenges of health literacy and accessing community services and support.[26] Prior research has shown that those from higher deprivation have significantly reduced healthy life years.[27] Reduced healthy life years are likely as a result of combined individual and geographical deprivation factors. At an individual level poor diet[28], poor housing[29] and an inability to afford travel to primary, secondary and tertiary care appointments.[30] There is also a suggestion of a lack of trust in public institutions, including the NHS, from the most deprived groups[31] – impacting their decision to access healthcare further. At a geographical level, there are fewer GPs per patient in more deprived areas and greater waits for non-urgent care.[32–33]

Our findings are suggestive of these challenges in our ILD population which raises important considerations for future policy, systems and research. For research, we need to understand the geographical and individual-level deprivation factors in more detail so that we can design a service that supports this group of patients. While this has been assessed more generally, revealing many layers of reason for delayed or challenging healthcare access (30), there may be specific challenges unaccounted for in ILD care given the significant unmet needs many in the service experience.[34] As a way to begin addressing these issues, we need to increase awareness and education of ILDs amongst patients, caregivers and primary care physicians working in such deprived areas. By going out into communities, into settings that such patients frequent, we can build this awareness but also trust. Policymakers must remain acutely aware of such groups in future service planning, especially in the context of rapid digital healthcare expansion, so that new services or interventions in ILD don’t inversely worsen inequalities for this group of patients. [35]

## Conclusion

From our dataset, we can cautiously infer that social deprivation impacts on health-seeking behaviours – with higher demands on acute healthcare services. However, once admitted in the context of an acute ILD-related admission, the association between deprivation and mortality outcomes becomes much more complex and multifactorial – and the impact of geographical deprivation reduced. Further studies are needed to examine this association in more detail, with careful consideration of geographical vs individual-level markers of deprivation.

**Figure 1:**
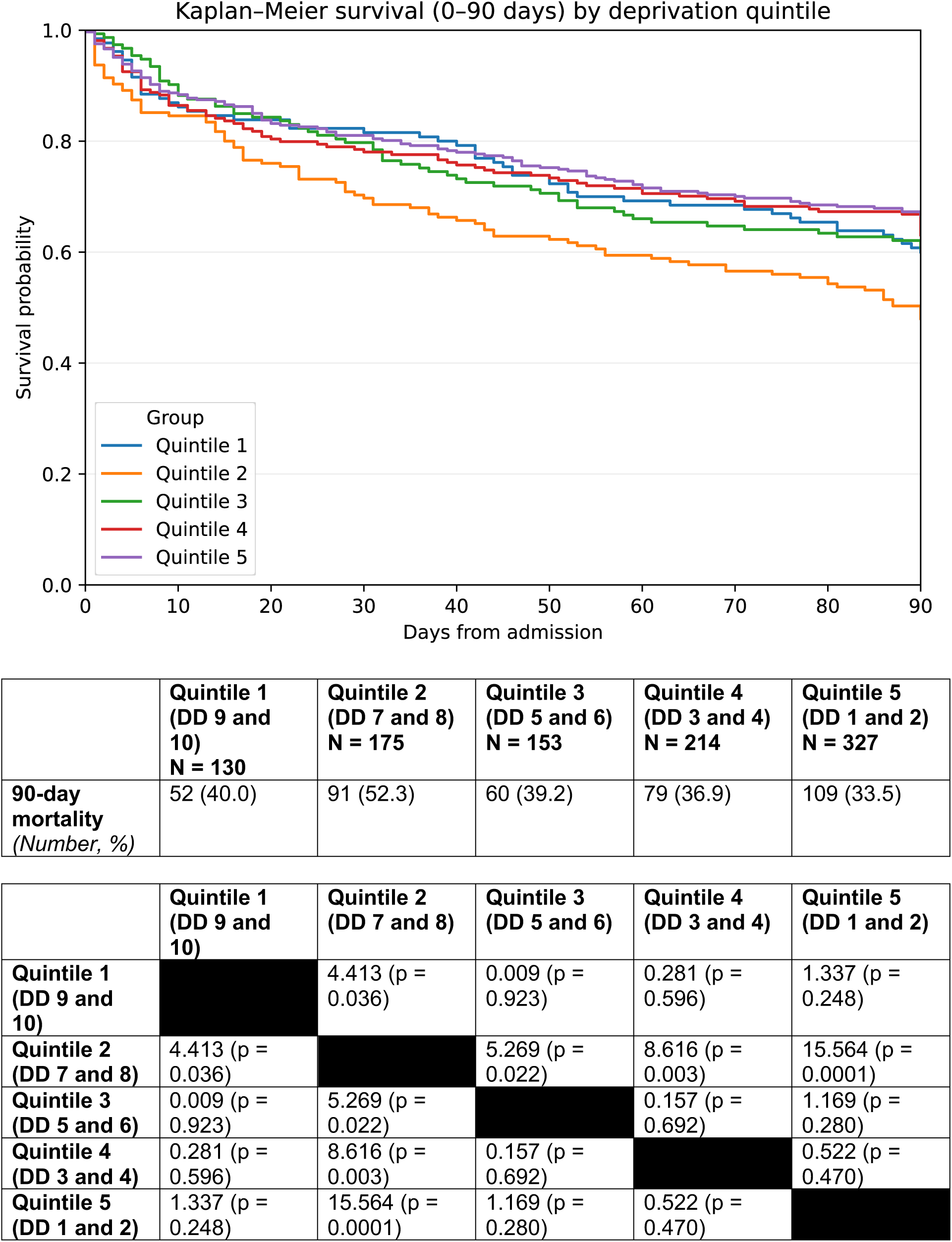
Kaplan Meier time-to-event analysis of 90-day all-cause mortality comparing deprivation quintiles, where quintile 1 represents the least deprived 20% and quintile 5 represents the most deprived 20%. The accompanying tables report 90-day all-cause mortality outcomes as number of events (death) and as a percentage of all ILD-related admissions within the relevant deprivation quintile, alongside log-rank pairwise comparisons of 90-day all-cause mortality between quintile groups. A p value of <0.05 was considered statistically significant. *Abbreviations: DD = deprivation decile*.

## Data Availability

The anonymised data supporting the findings of this study are available from the corresponding author upon reasonable request.

## Acknowledgements

Thank you to the Northern Care Alliance NHS Foundation Trust for providing sponsorship of the study.

Thank you to the National Institute of Health and Social Care Research for the Academic Clinical Fellowship programme which afforded LW protected time to conduct this study.

Thank you to Professor Andy Vail and Lancaster University Faculty of Health and Medical Sciences Maths and Statistics Hub for providing statistical guidance.

## Funding Statement

LW has funding from the NIHR as an Academic Clinical Fellow to fund 25% research time within her job plan. This fellowship did not provide any direct funding for this study.

The Northern Care Alliance NHS Foundation Trust provided sponsorship of the study. There was no monetary support attached to this.

## Notes

### Competing Interest Statement

The authors have declared no competing interest.

### Author Declarations

The study was approved by the Health Research Authority (reference 23/HRA/4562). Further approval from a Research Ethics Committee was not required due to use of retrospective secondary data. Secondary data was managed and used as per GDPR guidelines.

