## Supplementary Figure 1; Supplementary Table 1; Supplementary Table 2; Supplementary Table 3; Supplementary Table 4; Supplementary Table 5 for "Real world evidence of acute interstitial lung disease-related hospital admissions infers complex, multifactorial association between social deprivation and 90-day all-cause mortality outcomes: data from the North West of England"

### Supplementary Materials

### Figures

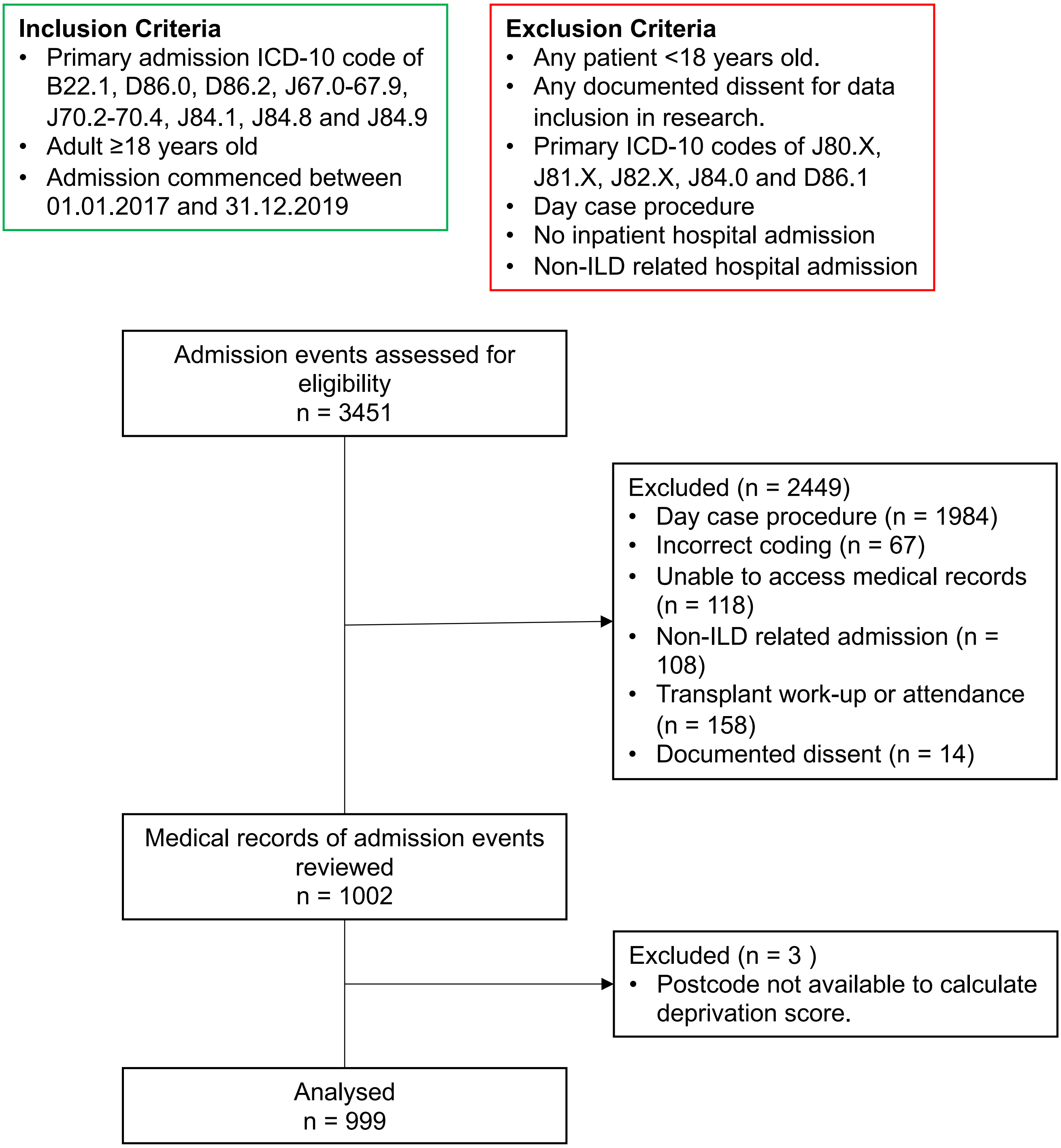


**Supplementary Figure 1:** Summary of inclusion and exclusion criteria, with flow diagram summarising case notes screened for inclusion and reasons for exclusion at each stage.
*Abbreviations: ICD-10 – international statistical classification of diseases and related health problems 10th revision; ILD – interstitial lung disease.*

### Tables

| **Age** | | | | | |
|---|---|---|---|---|---|
| | **Quintile 1** | **Quintile 2** | **Quintile 3** | **Quintile 4** | **Quintile 5** |
| **Quintile 1** | | 1.000 | 1.000 | 1.000 | **0.0006*** |
| **Quintile 2** | 1.000 | | 1.000 | 1.000 | **<0.0001*** |
| **Quintile 3** | 1.000 | 1.000 | | 1.000 | 0.161 |
| **Quintile 4** | 1.000 | 1.000 | 1.000 | | **0.003*** |
| **Quintile 5** | **0.0006*** | **<0.0001*** | 0.161 | **0.003*** | |
| **CCI** | | | | | |
| | **Quintile 1** | **Quintile 2** | **Quintile 3** | **Quintile 4** | **Quintile 5** |
| **Quintile 1** | | 1.000 | 1.000 | 1.000 | 0.226 |
| **Quintile 2** | 1.000 | | 1.000 | 1.000 | **0.016*** |
| **Quintile 3** | 1.000 | 1.000 | | 1.000 | 0.435 |
| **Quintile 4** | 1.000 | 1.000 | 1.000 | | **0.003*** |
| **Quintile 5** | 0.226 | **0.015*** | 0.435 | **0.003*** | |

**Supplementary Table 1:** Full summary of corrected P values from pairwise comparison of age and Charlson Comorbidity Index across quintiles using Dunn's test with subsequent Bonferroni correction.
Statistically significant values are marked in bold and with an asterixis (*).

*Abbreviations: CCI – Charlson Comorbidity Index*

| Smoking Status | | | | | | | | | | | | | | | |
|---|---|---|---|---|---|---|---|---|---|---|---|---|---|---|---|
| | Quintile 1 | | | Quintile 2 | | | Quintile 3 | | | Quintile 4 | | | Quintile 5 | | |
| | Residual | Raw | Corrected | Residual | Raw | Corrected | Residual | Raw | Corrected | Residual | Raw | Corrected | Residual | Raw | Corrected |
| **Current smoker** | -0.578 | 0.563 | 1.00 | -0.814 | 0.415 | 1.00 | -0.111 | 0.911 | 1.00 | -0.602 | 0.547 | 1.00 | 1.524 | 0.128 | 1.00 |
| **Ex-smoker** | 0.151 | 0.880 | 1.00 | 0964 | 0.335 | 1.00 | 0.147 | 0.883 | 1.00 | 0.520 | 0.603 | 1.00 | -1.321 | 0.186 | 1.00 |
| **Never smoker** | 1.274 | 0.203 | 1.00 | 0.576 | 0.565 | 1.00 | -0.148 | 0.882 | 1.00 | -1.574 | 0.116 | 1.00 | 0.151 | 0.880 | 1.00 |
| **Not available** | -1.651 | 0.099 | 1.00 | -1.951 | 0.051 | 1.00 | 0.009 | 0.993 | 1.00 | 1.514 | 0.130 | 1.00 | 1.237 | 0.216 | 1.00 |
| **ILD Subtype** | | | | | | | | | | | | | | | |
| | Quintile 1 | | | Quintile 2 | | | Quintile 3 | | | Quintile 4 | | | Quintile 5 | | |
| | Residual | Raw | Corrected | Residual | Raw | Corrected | Residual | Raw | Corrected | Residual | Raw | Corrected | Residual | Raw | Corrected |
| **Not known** | -0.558 | 0.577 | 1.00 | -1.079 | 0.280 | 1.00 | 1.197 | 0.231 | 1.00 | 0.341 | 0.733 | 1.00 | 0.047 | 0.963 | 1.00 |
| **IPF** | 0.755 | 0.450 | 1.00 | 1.885 | 0.059 | 1.00 | -1.210 | 0.226 | 1.00 | -1.253 | 0.210 | 1.00 | -0.014 | 0.989 | 1.00 |
| **NSIP** | 2.714 | **0.007** * | 0.366 | -1.956 | **0.050** * | 1.00 | 0.328 | 0.743 | 1.00 | -0.064 | 0.949 | 1.00 | -0.453 | 0.650 | 1.00 |
| **CTD-ILD** | 1.243 | 0.214 | 1.00 | -0.367 | 0.713 | 1.00 | -0.696 | 0.486 | 1.00 | -0.341 | 0.733 | 1.00 | 0.237 | 0.812 | 1.00 |
| **HP** | -0.006 | 0.995 | 1.00 | 1.501 | 0.133 | 1.00 | -0.231 | 0.817 | 1.00 | 0.617 | 0.537 | 1.00 | -1.435 | 0.151 | 1.00 |
| **Drug-related** | -0.509 | 0.610 | 1.00 | 2.112 | **0.035** * | 1.00 | 1.433 | 0.152 | 1.00 | -0.789 | 0.430 | 1.00 | -1.566 | 0.117 | 1.00 |
| **Industry-related** | 0.430 | 0.667 | 1.00 | -0.649 | 0.516 | 1.00 | -0.456 | 0.649 | 1.00 | 1.092 | 0.275 | 1.00 | -0.367 | 0.713 | 1.00 |
| **Sarcoidosis** | -0.337 | 0.736 | 1.00 | 0.494 | 0.621 | 1.00 | -1.588 | 0.112 | 1.00 | -0.816 | 0.415 | 1.00 | 1.597 | 0.110 | 1.00 |
| **PFD** | -1.453 | 0.146 | 1.00 | -0.907 | 0.365 | 1.00 | 1.884 | 0.060 | 1.00 | 0.671 | 0.502 | 1.00 | -0.252 | 0.801 | 1.00 |

| | | | | | | | | | | | | | | | |
|---|---|---|---|---|---|---|---|---|---|---|---|---|---|---|---|
| **Unclassifiable** | -1.943 | 0.052 | 1.00 | -0.479 | 0.632 | 1.00 | -1.633 | 0.102 | 1.00 | 3.125 | 0.002 | 0.098 | 0.165 | 0.869 | 1.00 |
| **Other** | -1.197 | 0.231 | 1.00 | -1.264 | 0.206 | 1.00 | -0.588 | 0.557 | 1.00 | -1.277 | 0.201 | 1.00 | 3.115 | **0.002** * | 0.101 |
| **Antifibrotics** | | | | | | | | | | | | | | | |
| | **Quintile 1** | | | **Quintile 2** | | | **Quintile 3** | | | **Quintile 4** | | | **Quintile 5** | | |
| | Residual | Raw | Corrected | Residual | Raw | Corrected | Residual | Raw | Corrected | Residual | Raw | Corrected | Residual | Raw | Corrected |
| **Nintedanib** | -1.448 | 0.148 | 1.00 | 1.952 | 0.051 | 1.00 | 1.785 | 0.074 | 1.00 | -1.540 | 0.124 | 1.00 | -0.490 | 0.624 | 1.00 |
| **Pirfenidone** | -1.361 | 0.173 | 1.00 | 1.924 | 0.054 | 1.00 | -2.053 | 0.040 | 0.802 | -0.515 | 0.606 | 1.00 | 1.272 | 0.203 | 1.00 |
| **Nintedanib + Pirfenidone** | -1.361 | 0.718 | 1.00 | 1.971 | **0.049** * | 0.975 | -0.391 | 0.696 | 1.00 | -0.463 | 0.643 | 1.00 | -0.572 | 0.567 | 1.00 |
| **None** | 0.750 | 0.453 | 1.00 | -1.086 | 0.277 | 1.00 | 0.159 | 0.874 | 1.00 | 0.536 | 0.592 | 1.00 | -0.221 | 0.825 | 1.00 |
| **AEILD Status** | | | | | | | | | | | | | | | |
| | **Quintile 1** | | | **Quintile 2** | | | **Quintile 3** | | | **Quintile 4** | | | **Quintile 5** | | |
| | Residual | Raw | Corrected | Residual | Raw | Corrected | Residual | Raw | Corrected | Residual | Raw | Corrected | Residual | Raw | Corrected |
| **AEILD** | 0.569 | 0.570 | 1.00 | -0.248 | 0.805 | 1.00 | -1.257 | 0.209 | 1.00 | 1.698 | 0.089 | 1.00 | -0.692 | 0.489 | 1.00 |
| **Other** | -0.713 | 0.476 | 1.00 | 0.527 | 0.598 | 1.00 | 2.096 | **0.036** * | 0.541 | -1.471 | 0.141 | 1.00 | -0.179 | 0.858 | 1.00 |
| **Insufficient information for AEILD status** | 0.233 | 0.816 | 1.00 | -0.660 | 0.509 | 1.00 | -1.853 | 0.064 | 0.958 | -1.002 | 0.316 | 1.00 | 2.414 | **0.016** * | 0.236 |

**Supplementary Table 2:** Full summary of post-hoc analysis of categorical variables using standardised residuals with Bonferroni correction. Reported values for each category through pairwise comparison are shown as residual values, raw P values and the corrected P value with the Bonferroni correction for each data point.

Statistically significant values are marked in bold and with an asterixis (*).

*Abbreviations: ILD – interstitial lung disease; IPF – idiopathic pulmonary fibrosis; NSIP – non-specific interstitial pneumonia; CTD-ILD – connective tissue disease interstitial lung disease; HP – hypersensitivity pneumonitis; PFD – pulmonary fibrosis as a diagnostic label; AEILD – acute exacerbation of interstitial lung disease*

| Data Point | Missing Count | Missing % |
|---|---|---|
| **Oxygen (L/min) at admission** | 679 | 68 |
| **TLCO (mmHg)** | 596 | 59.7 |
| **FVC (L)** | 387 | 38.7 |
| **CRP** | 98 | 9.8 |
| **Lymphocyte count** | 35 | 3.5 |
| **Monocyte count** | 34 | 3.4 |
| **WCC** | 27 | 2.7 |
| **Neutrophil count** | 27 | 2.7 |
| **Admission Length (days)** | 4 | 0.4 |

**Supplementary Table 3:** Summary of number of missing data points for continuous variables within the retrospective dataset.

*Abbreviations: L/min – litres per minute; TLCO – transfer factor of the lung for carbon dioxide; mmHg – millimetres of mercury; FVC – forced vital capacity; L – litres; CRP – c-reactive protein; WCC – white cell count (total).*

| **Oxygen (L/min) at admission** | P value | **FVC (L)** | P value | **TLCO (mmHg)** | P value |
|---|---|---|---|---|---|
| **Age** | 0.171 | **Age** | 0.295 | **Age** | **0.019*** |
| ***CCI** | 0.571 | **CCI** | 0.619 | **CCI** | **0.015*** |
| **Deprivation Decile** | **0.0002*** | **Deprivation Decile** | **0.002*** | **Deprivation Decile** | **0.001*** |
| **90-day Mortality Outcome** | 0.080 | **90-day Mortality Outcome** | 0.178 | **90-day Mortality Outcome** | 0.295 |

**Supplementary Table 4:** Summary of logistic regression modelling to assess pattern of data missingness. In a logistic regression model, missing data was compared with age, Charlson Comorbidity Index, deprivation decile values and 90-day mortality outcomes.
Statistically significant values are marked in bold and with an asterixis (*).
Statistically significant values suggest association between missing data and the variable. This suggests data was consistent with a missing-at-random (MAR) mechanism.

*Abbreviations: L/min – litres per minute; TLCO – transfer factor of the lung for carbon dioxide; mmHg – millimetres of mercury; FVC – forced vital capacity; L – litres; CRP – c-reactive protein; WCC – white cell count (total); CCI – Charlson Comorbidity Index.*

| | HR | Lower 95% CI | Upper 95% CI | P value |
|---|---|---|---|---|
| **Age** | 1.021 | 1.010 | 1.035 | **0.003*** |
| **Age x Deprivation Decile** | 0.999 | 0.996 | 1.001 | 0.258 |
| **Sex** | | | | |
| Female | Reference | Reference | Reference | Reference |
| Male | 1.505 | 1.233 | 1.837 | **<0.0001*** |
| **Ethnicity** | | | | |
| White | Reference | Reference | Reference | Reference |
| Asian | 0.890 | 0.637 | 1.272 | 0.550 |
| Black | 1.060 | 0.454 | 2.471 | 0.894 |
| Not stated | 0.833 | 0.575 | 1.210 | 0.336 |
| **CCI** | 1.034 | 0.967 | 1.106 | 0.323 |
| **Deprivation Decile** | 1.110 | 0.925 | 1.336 | 0.256 |
| **ILD Subtype** | | | | |
| Not stated | Reference | Reference | Reference | Reference |
| IPF | 1.282 | 0.918 | 1.790 | 0.145 |
| NSIP | 1.044 | 0.695 | 1.568 | 0.837 |
| CTD-ILD | 0.995 | 0.651 | 1.520 | 0.980 |
| HP | 0.948 | 0.653 | 1.376 | 0.780 |
| Drug-related | 0.638 | 0.362 | 1.125 | 0.120 |
| Industry-related | 1.186 | 0.620 | 2.268 | 0.606 |
| Sarcoidosis | 1.058 | 0.612 | 1.824 | 0.839 |
| PFD | 1.598 | 1.164 | 2.195 | **0.004*** |
| Unclassifiable | 1.062 | 0.578 | 1.954 | 0.845 |
| Other | 1.291 | 0.798 | 2.090 | 0.298 |
| **Oxygen** | | | | |
| None | Reference | Reference | Reference | Reference |
| Long-term | 2.826 | 2.150 | 3.713 | **<0.0001*** |
| Ambulatory | 2.021 | 1.449 | 2.820 | **<0.0001*** |
| **AEILD Status** | | | | |
| Insufficient information | Reference | Reference | Reference | Reference |
| AEILD | 1.342 | 0.952 | 1.893 | 0.093 |
| Other ILD-related admission | 1.472 | 1.059 | 2.046 | **0.022*** |
| **Neutrophils** | 1.027 | 0.995 | 1.061 | 0.103 |
| **Monocytes** | 1.063 | 0.981 | 1.106 | 0.323 |
| **CRP** | 0.998 | 0.996 | 0.999 | **0.035*** |

| | | | | |
|---|---|---|---|---|
| **FVC (L)** | 1.007 | 0.991 | 1.022 | 0.407 |
| **TLCO (mmHg)** | 0.888 | 0.839 | 0.941 | **<0.0001*** |
| **Oxygen (L/min) at admission** | 0.977 | 9.934 | 1.022 | 0.321 |

**Supplementary Table 5:** Full results of multivariate cox regression analysis of 90-day all-cause mortality associated with interstitial lung disease-related hospital admissions, using multiple imputation modelling for FVC, TLCO and oxygen required at admission (litres) and an age x deprivation term, to assess impact of age differences on model outcomes.
Statistically significant values are marked in bold and with an asterixis (*).

*Abbreviations: HR – hazard ratio; CI – confidence interval; ILD – interstitial lung disease; CCI – Charlson comorbidity index; IPF – idiopathic pulmonary fibrosis; NSIP – non-specific interstitial pneumonia; CTD-ILD – connective tissue disease interstitial lung disease; HP – hypersensitivity pneumonitis; PFD – pulmonary fibrosis as a diagnostic label; AEILD – acute exacerbation of interstitial lung disease; CRP – C-reactive protein; FVC – forced vital capacity; L – litres; TLCO – transfer factor of the lung for carbon monoxide; L/min – litres per minute.*
